# Decision-making in patients with ALS: experiences and implications for decision support

**DOI:** 10.64898/2026.04.22.26351518

**Authors:** Masako Nagase, Keiko Hino, Ayumi Sakamoto, Masae Seo

**Affiliations:** Faculty of Health Care and Nursing, Juntendo University, Urayasu 279-0023, Chiba, Japan

## Abstract

Patients with amyotrophic lateral sclerosis (ALS) face critical decisions regarding life-sustaining treatments, such as invasive mechanical ventilation and percutaneous endoscopic gastrostomy. Advance care planning and shared decision-making are standard supportive frameworks but they often fail to account for structural pressures like progressive decline, shifting patient values, and fear of becoming a ‘burden’ that may influence decision-making. This study explores how patients with ALS interpret ventilator and care options amid progressive physical decline, thereby reconsidering approaches to decision support. Using a qualitative descriptive design, the researcher (a nurse/sociologist) conducted 2–3 hour home interviews with five purposively sampled patients with ALS. Data, including eye-tracking-aided responses, were analysed via Sandelowski’s framework. Rigour was ensured through team-based triangulation, independent coding by two researchers, and a reflexive audit trail. Subjective narratives were prioritised without medical record cross-referencing to capture patients’ experiences. Four categories emerged: (1) Rewriting clinical prognosis into a ‘narrative of exploration’ via peer models, where meeting active ventilator users transformed future perceptions; (2) The conflict between securing care infrastructure and the ‘burden’ on family, which greatly influenced the will to survive; (3) Existential fluctuation, where patients’ intentions shifted with daily fulfilment and family events; and (4) Governance of the body via pre-emptive technology use and training carers as physical extensions. Findings showed decision-making was a multi-layered process redefining life’s meaning within social resources. This necessitate shifting from independent to relational autonomy, where agency relies on care infrastructure, not physical ability. Treatment choice is a dynamic exploration requiring ‘narrative companions’ to support existential fluctuations. Professionals must coordinate environments to reduce patient indebtedness. Limitations include the small, resource-advantaged sample (N = 5) and reliance on subjective narratives without medical record verification. Living with ALS means governing a new self through relational support and continuous dialogue.

## Introduction

In November 2019, the death of a patient with amyotrophic lateral sclerosis (ALS) by assisted suicide in Japan brought the profound difficulties of living with neurological diseases into sharp focus. However, societal debates surrounding euthanasia often overshadow the empirical exploration of how patients navigate the disease’s critical clinical trajectory. ALS is characterised by a progressive loss of motor function, leading to pivotal clinical decision points, including the initiation of non-invasive ventilation, tracheostomy with invasive mechanical ventilation, and percutaneous endoscopic gastrostomy [1]. In Japan, the care pathway involves multidisciplinary teams and specialised clinics, which can reduce emergency hospitalisations and improve survival rates [2]. Although medical insurance has covered home mechanical ventilation since the 1990s—allowing many patients to live at home for over 10 years—the decision-making process within these structural supports remains complex.

Decision-making in ALS extends beyond ‘medical choices’ such as ventilation, nutrition, and the care environment; it is a process of renewing the ‘meaning of life’ within relationships and the supportive environment [3]. Although advance care planning and shared decision-making are recommended, they have limitations, such as an over-reliance on documentation and the difficulty of predicting the future [4,5]. Additionally, the implementation of advance directives in the ALS field is not uniform [6]. Considering these gaps, this study aims to elucidate the experiential decision-making process through the narratives of patients with ALS.

In ALS, as the disease progresses, a series of critical decisions arise regarding non-invasive ventilation, invasive mechanical ventilation, gastrostomy, place of care, and communication support [1]. These are not isolated ‘decision events’ but should be viewed as a dialogue process that requires repeated consideration in response to changing conditions. Consequently, decision-making support must go beyond the mere presentation of medical information to include support that encompasses lifestyle, values, and relationships [7].

Advance care planning is internationally defined as a ‘process’ for sharing preferences regarding future treatment and care, documenting them as necessary, and reviewing them considering changing circumstances [5]. However, the institutionalisation of living wills (advance directives) tends to require ‘fixing one’s future intentions in the present’ and has been criticised for practical failures and side effects (patient burden, over-reliance on documentation) [4]. Even in systematic reviews and meta-analyses limited to ALS, the possession and creation of advance directives vary significantly across studies, and a downward trend has been observed in recent years, suggesting that advance care planning/advance directives is not a framework that ‘spreads naturally’ [6].

Decision-making regarding ALS needs to be viewed not as an individual’s rational choice but as ‘relational autonomy’ formed within the interdependence of the patient, family, and healthcare professionals [3]. Qualitative research on multidisciplinary care indicates that decision-making is shaped by structure (institutions and resources), interaction (team relationships), and individual factors—highlighting decision-making as the ‘design of life infrastructure’ [6]. Recent qualitative research indicates that end-of-life decision-making is linked to coping and oscillates between avoidance and preparation [8]; that in cases such as gastrostomy, the ability to ‘decide at one’s own pace’ supports a sense of control more than the mere ‘presence or absence of choice’ [9]; and that decision-making regarding home mechanical ventilation is diverse and changes over time [10]. However, the details of decision-making experiences under conditions of severe paralysis and significant communication constraints (e.g. eye-tracking devices) have not been sufficiently described, and there is a lack of insights necessary for the redesign of support [11].

This study explores the decision-making process by which patients with ALS select their treatment and care environments as an interaction among relationships, social resources, and the communication environment, and to elucidate its meaning through patients’ narratives.

### Aim

The purpose of this study is to clarify the experiences through which patients with ALS make decisions when selecting treatments and care environments, and to rethink approaches to decision support that go beyond the frameworks of advance care planning and shared decision-making.

## Materials and methods

### Design

This study adopted a qualitative descriptive research design. This approach was chosen to gain a comprehensive description of patients’ narratives within their ‘lifeworld’ without the necessity of developing a formal theory, aligning with our exploratory aim to capture decision-making as an experiential and relational process.

### Participants

We used a purposive sampling method to recruit patients diagnosed with ALS or motor neurone disease (MND) who were already exhibiting symptoms within the ALS spectrum, all of whom had faced decisions regarding ventilators or their care environment. Participants were recruited through personal contacts. The recruitment period for research participants was set from 17 April 2019 to 28 February 2020. Although the sample size was small (N = 5), we aimed to obtain detailed and informative accounts of their experiences.

### Data collection

Semi-structured interviews were used for data collection. Participants were asked to discuss the following points:

- Experiences of selecting treatment and care environments; for example, ‘Could you tell me specifically about the discussions held regarding the use of a ventilator?’ ‘When and how was the decision made to add a gastrostomy tube?’ ‘Why was the decision made to live at home rather than in a care facility?’
- Conflicts arising between ‘the present’ and ‘the future’.
- Interactions with healthcare professionals and family.
- Events and values that influenced decision-making.

Interviews were conducted face-to-face, and audio recordings were transcribed verbatim to create the data. Where participants had provided files summarising their responses after being informed of the questions in advance, these were also used as data.

The interviews were conducted by the author herself at the research participants’ homes. The author is a nurse and a researcher in nursing science and medical sociology. Although prior contact had been made with the research participants via email, the interview was the first in-person meeting. Each interview lasted between two and three hours, with breaks for refreshments and casual conversation. Neither family members nor healthcare professionals were present during the interviews, although family members or specialist care providers were available in an adjacent room. This study utilised only the interview data and profiles compiled by the participants themselves; no cross-referencing with medical records was undertaken.

## Data analysis and audit trail

Data were analysed using qualitative descriptive analysis, following the methodological framework proposed by Sandelowski [12]. This approach was chosen to provide a comprehensive summary of participants’ experiences in everyday terms, staying close to the data with low-inference interpretation.

To ensure a rigorous audit trail, the following multi-step process was employed: Unit of analysis: The unit of analysis was defined as meaning units (sentences or paragraphs) related to the research questions.

Coding process: Initial open coding was conducted to capture the manifest content. These codes were then compared and grouped into sub-categories and categories through the constant comparative method.

Team-based triangulation: To mitigate interpretive bias, two researchers independently coded the first two transcripts. They then met to compare their findings and develop a preliminary codebook. Any disagreements were resolved through consensus or consultation with a third researcher (a specialist in qualitative research). This iterative process ensured the reliability of the categorisation.

Reflexivity: The researchers maintained a reflexive journal throughout the analysis to acknowledge their positionality as nurses/researchers/medical sociologist and to prevent personal clinical biases from overshadowing the participants’ narratives.

### Ethical considerations

This study was conducted with the approval of the Ethics Review Committee of the affiliated institution (Junkanrin No. 19-9; date of approval: 22 April 2019). Participants were informed about the research purpose, anonymity, voluntary participation, and the freedom to withdraw at any time, and written consent was obtained.

## Results

Five research participants were involved. Two of these (Cases B and C) were already fitted with ventilators and, due to quadriplegia, interviews were conducted via eye-tracking communication devices. One participant (Case D) underwent non-invasive positive pressure ventilation therapy only at night.

Analysis revealed that decision-making for patients with ALS is not merely about ‘choosing treatments’, but rather a dynamic, multi-layered process of re-weaving the ‘meaning of life’ within social resources and relationships with others. The four categories are detailed below. (See Table 1)

**Table 1.**
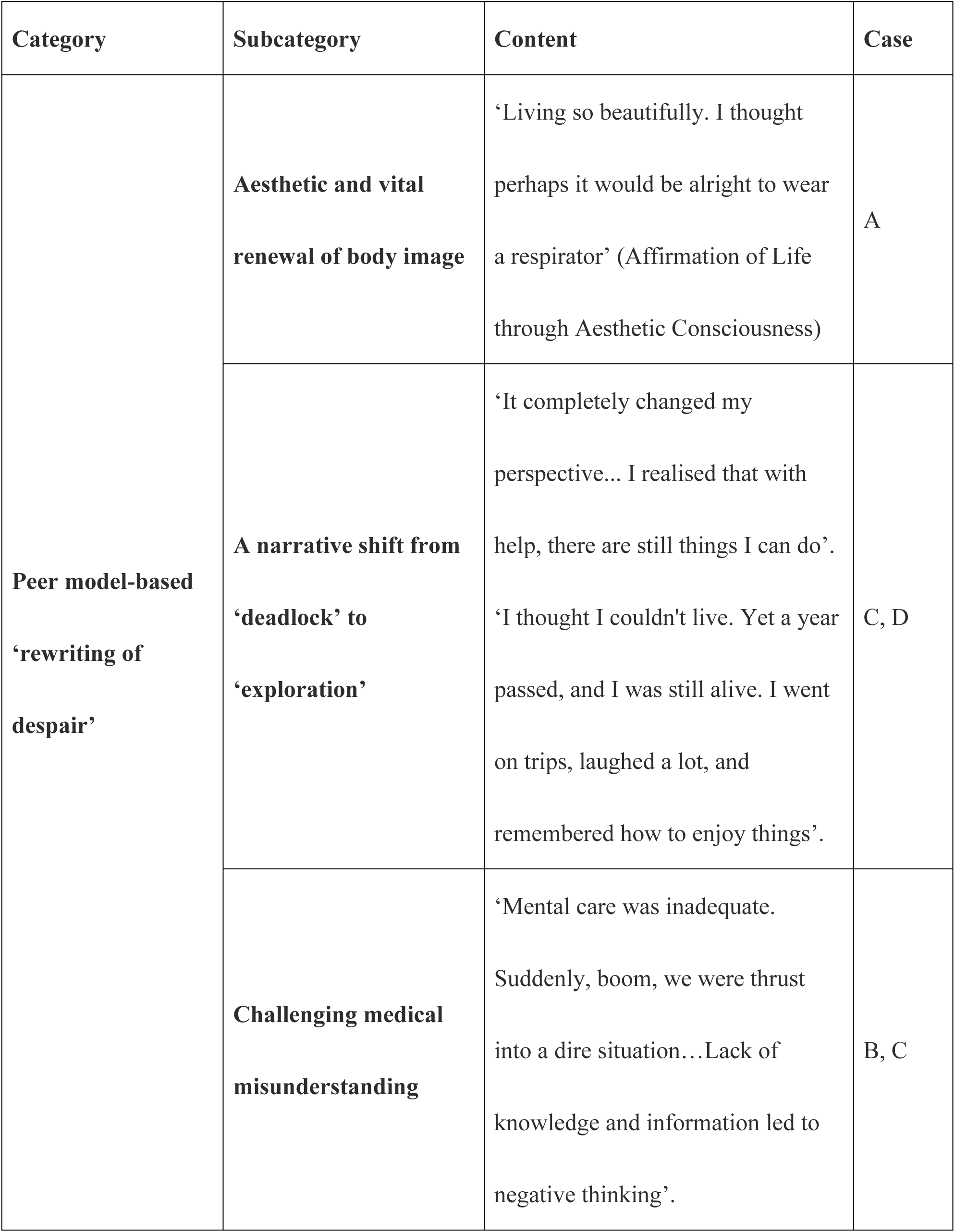
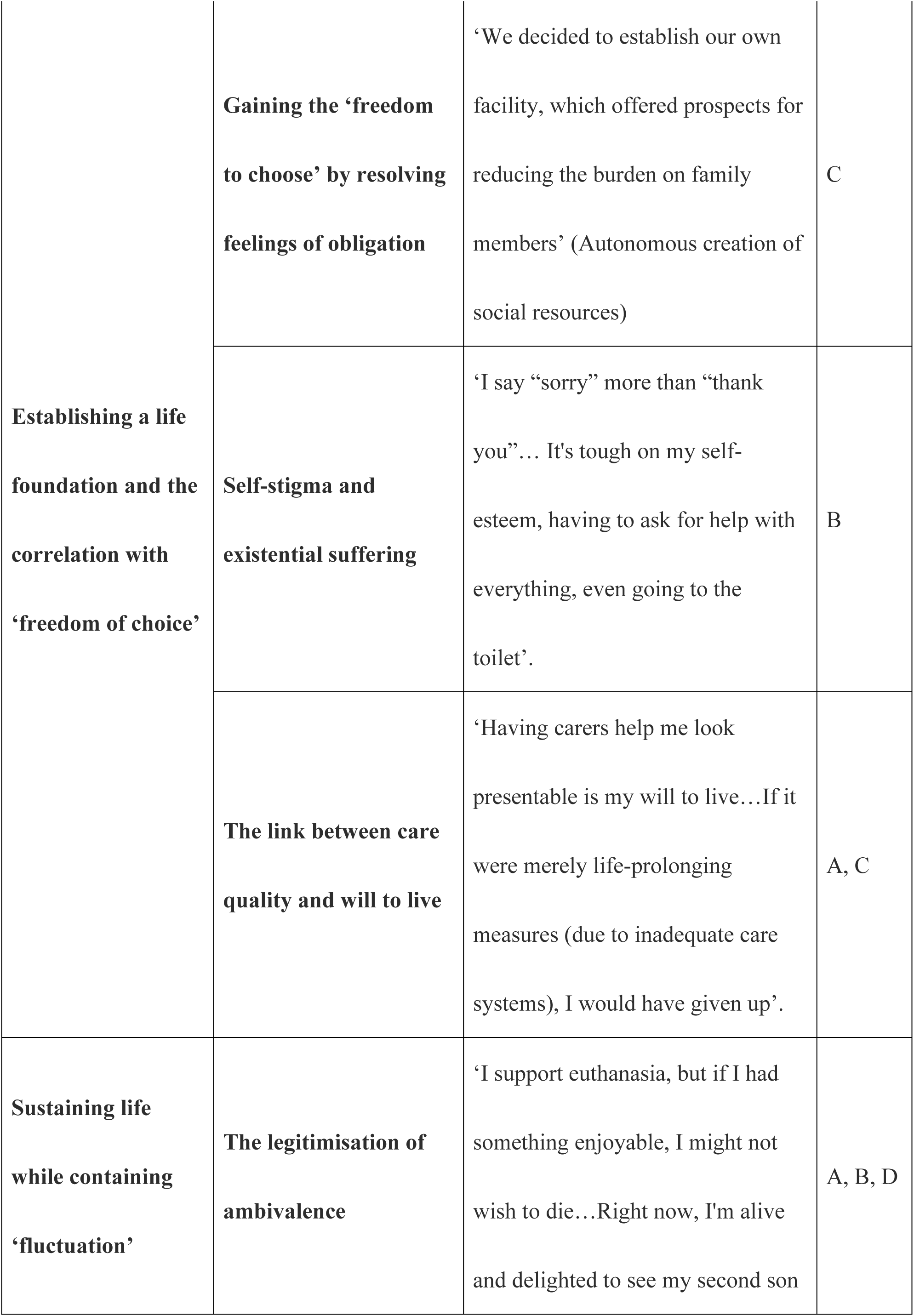
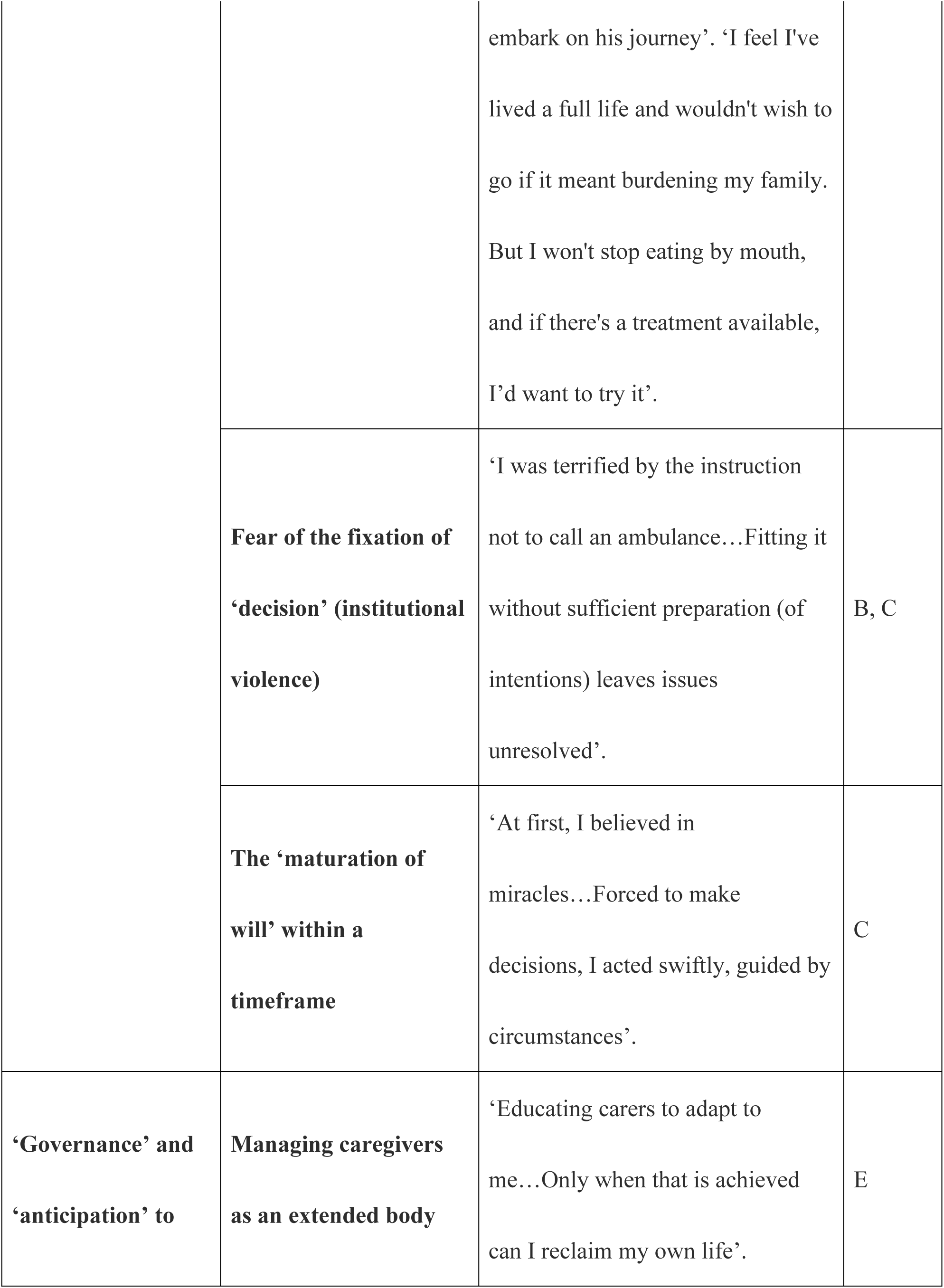
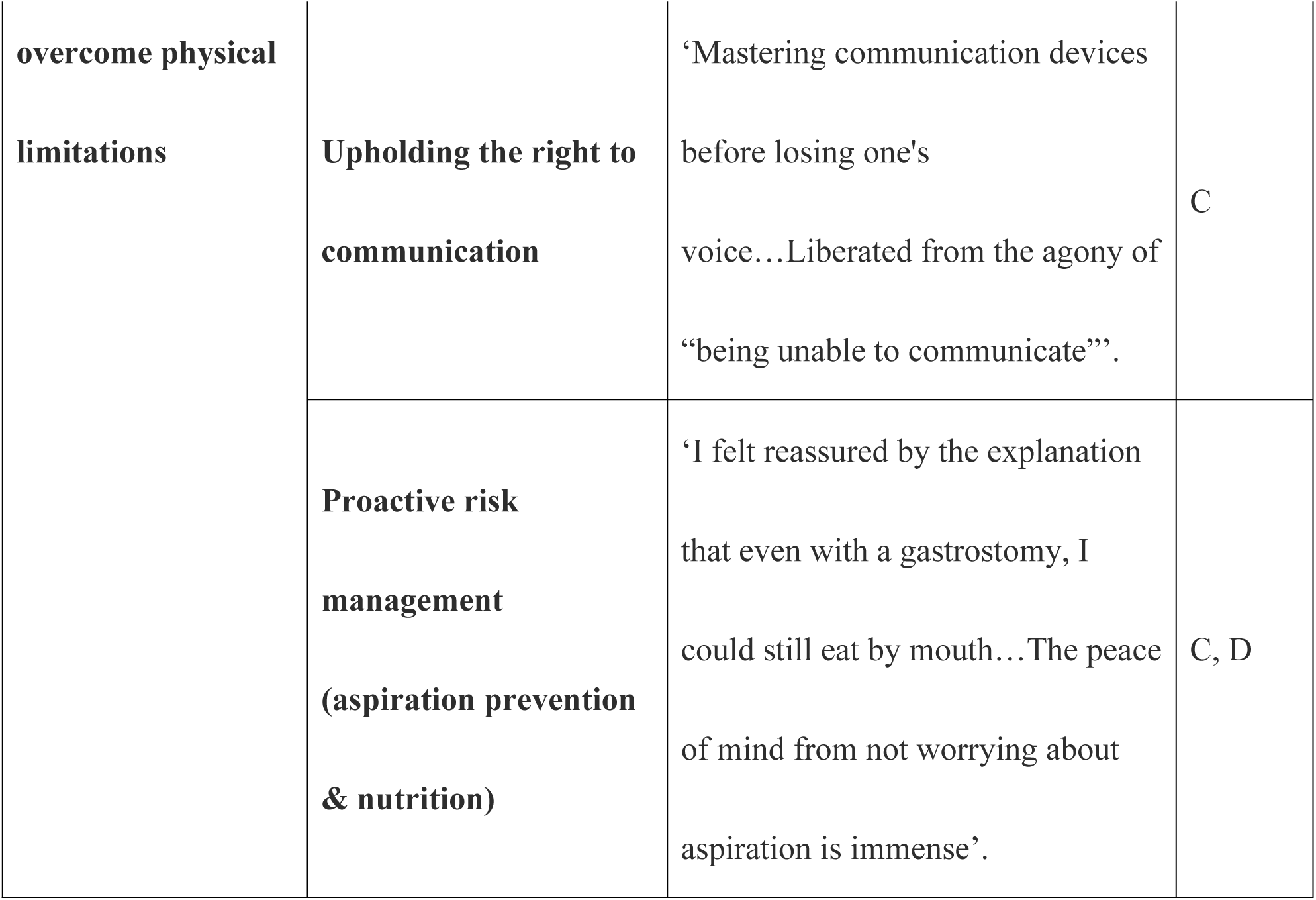
Characteristics of Decision-making Experienced by Patients with ALS

### Rewriting clinical prognosis: transition to a ‘narrative of exploration’ via peer models

Immediately after diagnosis, patients are confined within the medical information framework provided by healthcare professionals, which focuses on ‘functional loss’, leading them to perceive the future as ‘chaos’. However, encountering peers (fellow patients) who remain active while using ventilators acts as a catalyst, reconstructing the medical ‘prognosis’ into a concrete ‘continuation of life’.

> *‘I had assumed I would be confined to bed and unable to do anything. But when a nurse invited me to meet a fellow patient living at home with a ventilator, I saw them actively going out with a carer’s help and living life with strength. It completely changed my perspective.’ (Case C)*

> *‘(Seeing Case E) They live so beautifully. I began to think wearing a ventilator might be acceptable. Initially, I didn’t even know the difference between ALS and muscular dystrophy, but seeing them became a source of support.‘ (Case A)*Here, the ‘aesthetic awareness’ and ‘activity’ demonstrated by peers present a ‘quality of life’ not discussed within the medical context, becoming the starting point for patients to rewrite their own narratives from ‘passive victims’ to ’active explorers’.

### The foundation of relational autonomy: infrastructure development and the conflict with ‘burden’

Freedom of decision-making is determined not only by an individual’s internal will but also by structural factors such as the availability of care resources. Choices made in the absence of adequate care resources carry the risk of being forced into abandonment driven by a ‘burden on others’ and lacking true autonomy.

> *‘The reality is existing home care agencies are stretched thin and turning people away… We had resigned ourselves, thinking the device was merely a life-prolonging measure that would only increase the family’s burden…(Afterwards) we decided to set up our own agency ourselves, and the prospect of reduced burden became visible (which led to our decision).’ (Case C)*

> *‘My self-respect, asking for help with everything from toileting… It’s hard to ask my son or husband. “Sorry” has become more frequent than “thank you”’. (Case B)*The ‘self-initiated establishment of a care service’ seen in Case C represents a process of securing autonomy by designing one’s own solution to insufficient social resources. Conversely, the phrase ‘I’m sorry’ in Case B symbolises the pain of care resource shortages, which transforms the patient’s existence into a ‘liability’.

### The right to existential fluctuation: resistance to fixed intentions

An ALS patient’s will to survive is constantly fluid, influenced by physical condition and the quality of care. It is essential not to view this ‘fluctuation’ as indecisiveness but to accept it as a natural process of life. Pressure from the medical side to make an early ‘final decision’ inflicts severe existential suffering on the patient.

> *‘I’m in favour of euthanasia…But if I had something enjoyable happening or a goal at that time, I might not want it. So I don’t know.’ (Case B)*

> *‘I’d decided from the start not to have a ventilator…But now I’m alive and delighted at my second son’s new beginnings. If I want to live, shouldn’t I be able to live whenever I choose? That “do not call an ambulance” directive made me tremble’ (Cases A and B)*Treating a single expression of intent as a ‘fixed instruction (advance directive)’ exposes a structural failure that halts the ‘process of living’ the patient continually generates.

### Governance of the body and pre-emptive action: technology and the ‘selfification’ of others

During the process of losing bodily functions, patients attempt to maintain existential agency by acquiring skills before functional loss (pre-emptive action) and by training carers as extensions of their own bodies.

> *‘I train carers to adapt to me…Only then can I reclaim my life’ (Case E)*

> *‘Before losing my voice, I mastered operating the communication device…liberating myself from the agony of “being unable to communicate”’. (Case C)*This constitutes an endeavour to recover subjectivity as a ‘living person’ beyond the role of ‘patient’ by continuing to govern one’s own lifeworld, even in a state of total dependency.

## Considerations

The data from this study suggest that decision-making among people with ALS is not merely a matter of choosing a treatment or deciding on life-sustaining measures; rather, it is a dynamic and multi-layered process in which the meaning of ‘how to live’ is constantly redefined within the context of social resources, relationships with others, changes in physical function, and future prospects. Furthermore, decision-making was viewed not as a process concluded by an individual’s internal preferences but as a practice embedded in concrete experiences such as encounters with peers, the availability of care resources, shifts in intentions, and the acquisition of communication skills. These findings highlight the need to rethink and promote shared decision-making and advance care planning in patients with ALS not merely as activities to confirm and document preferences at an early stage, but as interactive processes that support patients in continuously updating their self-image of the future [4–6].

### The meaning of autonomy for ALS Patients: from independent autonomy to ‘relational autonomy’

This study revealed that, in the decision-making of patients with ALS, clinical prognosis is not perceived as fixed but is rewritten through encounters with others. Case C stated that immediately after receiving the diagnosis, they thought, ‘I will be bedridden and unable to do anything’; however, their perspective changed completely after meeting a fellow patient living at home on a ventilator, who even went out. The use of a ventilator is not merely a matter of ‘prolonging life’; it is a device that is essential for living life to the full. Although they find themselves in a situation where they must rely on carers for all aspects of daily living, and are aware that they may eventually become totally locked-in syndrome, they have not lost hope in life. This represents a paradigm shift from the conventional understanding of a ventilator as a means of ‘life-prolongation’ to one of a ‘life-support device’. Case A also stated that, upon encountering a patient who was ‘living a tidy life’ while using a ventilator, they thought, ‘Perhaps it would be okay for me to use a ventilator too’. What is significant here is that patients did not merely acquire new medical information; rather, through the concrete examples of others, they discovered aspects of the sustainability of daily life and quality of life that could not be fully captured by the ‘prognosis of functional loss’ provided by healthcare professionals. Peers served not only as sources of information but also as mediators in reconstructing a future self-image. This resonates with the argument that future-oriented decision-making in ALS is not a rational choice made by an isolated individual but rather an endeavour to envision ‘who one might become’ within a network of relationships [3]. This is also consistent with the model proposed by Hogden et al. [13], which demonstrated that decision-making in ALS is influenced not only by individual factors but also by support systems and the quality of interactions. Consequently, the current findings suggest that prognosis explanations should not be limited to presenting ‘lost functions’ but must concretely demonstrate what kind of life can be reconstructed while receiving support. In particular, as the narratives of Cases A and C demonstrate, the ‘sense of beauty’ and ‘activity’ embodied by peers are crucial elements that are difficult to capture through clinical indicators, yet are deeply involved in patients’ decision-making. Given recent reports on the usefulness of online peer support for ALS, such connections with peers should not be positioned as supplementary support but as a core resource for decision-making support [14].

Furthermore, this study specifically demonstrated that freedom of decision-making does not depend solely on the individual patient’s will but is largely determined by the availability of care infrastructure and support resources. In Case C, as existing home care agencies were overwhelmed and unable to provide support, the patient initially gave up on the idea of using a ventilator, believing that ‘its use is merely a life-prolonging measure that only increases the burden on the family’. However, after the prospect of reducing this burden emerged through the practical step of ‘setting up an agency themselves’, the decision became possible. This signifies that the patient was not passively choosing from existing resources but rather reconstructing the conditions for autonomy by reshaping social resources. Conversely, the statement in Case B—‘I say “I am sorry” more often than “thank you”—vividly illustrates the distress experienced when, due to a lack of support, dependence is perceived not as gratitude but as a debt. Choices regarding treatment and care were not determined solely by physical condition but were effectively constrained by emotions such as ‘I cannot rely on my family any further’ and ‘I do not want to impose a burden on them’. This is consistent with the perspective of ‘relational autonomy’, which views autonomy not as an internal capacity but as something supported by relational and social conditions [15]. Furthermore, a series of studies by Oliver and colleagues have highlighted that decision-making regarding ventilation and nutritional management in ALS/MND is strongly influenced not only by the progression of the disease itself but also by the support system and the availability of services [16–18]. The current findings indicate that it is insufficient to simply ask patients with ALS ‘what they want’; one must simultaneously ask ‘what choices are available to them’. Consequently, to make shared decision-making meaningful, it is necessary to position the development of infrastructure—such as home nursing, care staff, respite care, access to support schemes, and communication support—as an integral part of decision-making support, alongside explanations of treatment options.

### Reconstructing the narrative of treatment choice: from clinical prognosis to a ‘narrative of exploration’

This study presented a perspective that views the fluctuation of patients’ wishes over time not as ‘indecision’ or a ‘lack of consistency’, but precisely as the ‘process of continuing to embrace life’ itself. Case B stated: ‘I am in favour of euthanasia. However, if I have something enjoyable or a goal at that moment, I might not want to go through with it. So, I don’t know.’ This testimony candidly expresses that one’s intentions regarding death do not exist as fixed beliefs, but can change depending on one’s hopes, relationships and the meaning of life at that particular moment. Furthermore, as seen in the testimonies of Cases A and B, experiences such as ‘I had decided from the outset not to use a ventilator, yet here I am alive and delighted to see my son’s coming-of-age ceremony’, or the sensation of ‘shuddering at the instruction not to call an ambulance’, demonstrate that a will once expressed can be reinterpreted in the light of subsequent life experiences. What emerges from this is not that patients’ wishes are immature, but rather that, given the fluid nature of life itself, it is only natural for wishes to change. This finding is consistent with the view that advance care planning is an ongoing process [5]. Concurrently, it relates to the critique of over-reliance on fixed documents such as living wills and advance directives [4]. Furthermore, research on end-of-life care for patients with ALS/MND and those using home ventilators has reported that patients’ and their families’ decision-making changes over time, oscillating between avoidance and preparation [10]. Therefore, in clinical practice, rather than pressuring patients to make a ‘final decision’ at an early stage, it is considered necessary to value the patient’s freedom to change their mind and to continue exploring life, including treatment choices. A patient’s hesitation is, in essence, a ‘narrative of exploration’.

The loss of physical function does not necessarily imply a loss of agency. Rather, patients attempt to maintain control over their own lives by ‘anticipating’ and reconstructing their relationships with technology and others. Case E stated, ‘I instruct my carers to adapt to me. Only when they are able to do so can I reclaim my life.’ Here, carers are positioned not merely as supporters, but as an ‘extended body’ that realises the patient’s intentions within the concrete realities of daily life. Furthermore, Case C stated that by learning to operate communication devices before losing their voice, they were freed from the anguish of ‘being unable to communicate’. These testimonies demonstrate that even as severe disabilities progress, patients do not remain passive recipients of care but continue to strive to govern their own lifeworlds. Agency is not redefined by the scope of what one can do ‘on one’s own’, but rather by the ability to maintain one’s own pace and style while integrating appropriate technology, others and the environment into one’s life. This is consistent with the findings of van Eenennaam et al. [9]. They argue that, in decision-making regarding gastrostomy, the ability to ‘decide at one’s own pace’ supports a sense of control more than the mere ‘availability of options’. Furthermore, as voice banking research suggests, the early adoption of technology is not merely a preparatory measure but is also linked to the maintenance of self-identity [19]. This study also revealed that acquiring such technology is not merely a matter of securing a means of communication but is a practice that enables patients to continue being ‘people living their own lives’ without being confined to the role of ‘patient’. Consequently, support regarding percutaneous endoscopic gastrostomy, non-invasive ventilation, TIV and similar technologies should not be limited to the moment of deciding ‘whether to introduce them’; rather, it must be presented as an ongoing preparatory process designed to enable patients to express their own will and maintain control over their lives into the future.

## Discussion

As outlined above, this study portrays decision-making by patients with ALS not as a one-off choice of treatment but as a process of continually renewing the ‘meaning of life’ through the reinterpretation of prognosis, reorganisation of resources, acceptance of fluctuating intentions, and proactive management of the body and environment. This indicates the need to shift the focus of decision-making support in ALS from ‘making the patient decide’ to ‘creating the conditions for the patient to continue making decisions’. What is important is not to elicit a fixed answer from the patient at an early stage but to support the patient so that they can live while rewriting their own narrative time and again through encounters with peers, resource coordination by a multidisciplinary team, iterative dialogue that allows for changes in intentions, and the early securing of means of communication. In this sense, this study can be said to reinforce previous research—which noted that ‘difficult decision-making’ in ALS/MND requires careful advance planning and continuous support—through the concrete narratives of Japanese patients, and to present a perspective that reinterprets decision-making support in a more relational and practical manner [16,17,20].

### Study limitations

Although this study explored the decision-making processes of patients with ALS in depth through qualitative interviews, it is subject to several methodological limitations. First, the sample size was small (N = 5) and consisted mainly of patients who volunteered to participate from a specialist clinic; consequently, there are limitations regarding selection bias and the generalisability of the results [8,21]. In particular, data on patients on ventilators or those with communication difficulties are scarce, reflecting the challenge of data scarcity observed both domestically and internationally [1,22]. Second, interviews conducted using communication aids may have made it difficult to capture subtle nuances, and recording errors or mishearings may have occurred. Furthermore, the interviewer’s questioning style and respondent fatigue may have introduced recall bias or social desirability bias [4,5]. Third, a limitation is that changes following diagnosis could not be tracked from the time of study participation. In contrast, prospective long-term studies have clarified the impact of progressive communication impairment on healthcare involvement [21], and similar follow-up is desirable in this study. Regarding ethical considerations, obtaining consent via augmentative and alternative communication and using proxy respondents may pose challenges related to data accuracy and privacy [3,5].

The following four measures are proposed for future research:

- Surveys using larger samples that include multiple centres and diverse backgrounds.
- Follow-up studies targeting patients and families, along with multifaceted evaluations using mixed-methods research.
- Standardisation of data collection using augmentative and alternative communication-compatible interview manuals and online tools.
- Addressing practical challenges through research designs involving user-participatory research (user-centred design).

The limitations outlined above suggest that the current findings may be dependent on specific situations and contexts. When interpreting the results, it is necessary to consider sample bias and the constraints of data collection methods, as well as to conduct further verification in other cultural and institutional settings.

## Conclusion

This study demonstrated that decision-making for people with ALS is a dynamic process that goes beyond treatment choices, involving the re-weaving of ‘the meaning of life’ within the context of relationships, social resources, and physical changes. Encounters with peers encouraged the reconstruction of future visions; the availability of support resources influenced the scope of choice; and fluctuations in intentions should be understood as natural changes accompanying the process of living. Furthermore, proactively arranging communication technologies and care relationships is crucial for maintaining autonomy. Consequently, decision-making support for patients with ALS requires an approach that focuses not on fixed confirmation of intent but on creating conditions that enable patients to continue making decisions through iterative dialogue, resource coordination, peer support, and communication assistance.

This study is a small-scale case study, and its findings are limited. However, it presented a form of ‘autonomy’ enabling patients with ALS to maintain control and dignity. Living with ALS is not a continuous series of losses but a process of ‘governing a new self’ based on relationships with others.

## Data Availability

The data used in this study contain information that could identify the participants. For this reason, we have anonymised it and included references to parts of it within the paper. Data requests may be sent to Research Ethics Committee. Faculty of Health Care and Nursing, Juntendo University. 2-5-1, Takasu, Urayasu, Chiba 2790023

## Acknowledgments

We are deeply grateful to the five research participants who, despite facing communication difficulties, kindly agreed to take part in this study. We would like to thank Editage (www.editage.jp) for English language editing.

